# Immunometabolic dysregulation in depression predates illness onset and associates with lower brain gray matter volume

**DOI:** 10.1101/2025.08.14.25333723

**Authors:** Ye Ella Tian, Corey Giles, Maria A. Di Biase, Robin Cash, Vanessa Cropley, Andrew Zalesky

## Abstract

Depression often co-occurs with physical health conditions, including heart disease, diabetes and obesity. While dysregulation of the immunometabolic system is posited to underpin several of these comorbidities, the course of immunometabolic dysregulation in depression and its impact on structural brain changes linked to the disorder remain poorly understood. Using brain imaging and metabolomics data from the UK Biobank, we comprehensively evaluated cross-sectional and longitudinal immunometabolic profiles in depression, including inflammatory markers, lipoprotein lipids, fatty acids, amino acids, glycolysis metabolites and various low-molecular-weight metabolites. We found that depression is characterized by a relatively persistent pattern over time of elevated systemic inflammation, upregulated very-low-density lipoprotein and lipids and downregulated high-density lipoprotein with small-to-modest effect sizes (|Cohen’s d| = 0.01-0.16), and predates illness onset (mean prodromal period: 7 years). We mapped network-level systemic changes in metabolites, implicating the core role of glycolysis in depression-related metabolic dysregulation. We also showed that peripheral immunometabolic dysfunction, particularly elevated inflammation, is associated with lower brain gray matter volume in depression. We concluded that altered lipids and inflammatory markers predate the onset of depression, remain altered throughout the illness course and associate with lower gray matter volume. By comprehensively profiling immunometabolic dysfunction in depression and related brain changes, our work highlights the importance of monitoring and managing chronic low-grade inflammation and altered lipid and glucose metabolism in the disorder.

## Introduction

Major depressive disorder is a highly prevalent mental health condition, with an approximate 12-month prevalence of 6% on average and affecting about 20% of the global population at some point in their lifetime ^1^. Depression is associated with a heightened risk of cardiometabolic conditions, including coronary heart disease ^2^, stroke ^3^, obesity ^4^ and diabetes mellitus ^5^. Emerging evidence suggests that chronic physical illness comorbidity and somatic symptoms in depression may be linked with immunometabolic dysregulation, encompassing chronic low-grade inflammation, oxidative stress and imbalanced energy homeostasis ^6,7^. Indeed, meta-analyses ^8–10^ and studies using high-throughput metabolomics ^11–14^ have identified associations between depression and dysregulation of peripheral metabolic biomarkers including lipid metabolism. It was found that depression associates with downregulation of high-density lipoprotein (HDL) and upregulation of very-low-density lipoprotein (VLDL) and triglycerides as well as alterations in a variety of plasma lipids, fatty acids and low-molecular-weight metabolites.

However, few studies ^15,16^ have examined whether immunometabolic dysregulation predates the onset of depression. If this dysregulation is prodromal, we can: i) hypothesize that immune-metabolic disturbances contribute to disease development, ii) identify high-risk individuals before illness onset, and iii) facilitate early detection of subclinical depression.

Prospective studies are thus required to understand whether immunometabolic dysregulation predates depression onset and confers increased risk of depression.

Immunometabolic dysfunction may also be a consequence of depression, due to long-term illness, behavioral changes and other factors. Individuals with depression are more likely to engage in unhealthy habits, such as smoking, excessive alcohol consumption, poor diet and physical inactivity ^7,17^, and are less likely to comply with medical advice ^18^. Chronic stress and associated behavioral changes related to depression can also lead to dysregulation of the hypothalamic-pituitary-adrenal axis and inflammation, leading to dyslipidemia ^19^ and increased risk of cardiometabolic diseases ^20^. Although antidepressant (e.g., selective serotonin reuptake inhibitors) side effects can disturb metabolic homeostasis ^21^, the effect of antidepressant pharmacotherapies on metabolic function remains inconclusive ^22,23^.

Longitudinal assessments are required to understand whether immunometabolic dysregulation is an inherent trait related to depression pathophysiology or reflects a state-dependent clinical characteristic of the disorder. A trait-like persistent dysregulation over the course of illness would suggest the need for treatment targeting the dysregulation.

Most current immunometabolic studies in depression focus on a single metabolic marker, overlooking the complex interrelationships between metabolites. To overcome this limitation, metabolism can be conceptualized as chains of enzyme-catalysed reactions that can be represented as a network ^24^. Connections in such a network can be drawn between pairs of metabolites, where up-or down-regulation of one metabolite in the pair leads to a similar effect in the other metabolite. Network-analysis may thus facilitate identification of systemic metabolic changes in depression that may not be detectable when relying solely on individual metabolite levels ^25,26^. In this study, we thus adopt network models to study systemic metabolic changes.

Depression is linked with structural and functional brain changes across multiple cortical and subcortical regions ^27–30^. However, the influence of peripheral immunometabolic dysfunction on these brain changes remains inconclusive ^31–33^. Although most peripheral lipids and lipoproteins cannot directly access the brain due to the blood-brain barrier (BBB) ^34,35^, emerging evidence suggests that metabolic disorders, including diabetes mellitus, dyslipidemia and obesity, are associated with brain gray matter atrophy and functional brain changes ^36–38^, possibly explained by neuroinflammation induced by peripheral VLDL and cytokines ^39^. For example, peripheral inflammatory cytokines can initiate central inflammatory responses by signalling through the BBB ^40^, passing through the circumventricular organs ^41^ or via binding to the vagus nerve ^42,43^. Further work is needed to understand how peripheral metabolic dysfunction and inflammation potentially impacts brain structure and function.

Here, we curate longitudinal peripheral immunometabolic profiles from individuals with depression using a comprehensive panel of circulating inflammatory and metabolomic markers from the UK Biobank ^12,44,45^. We evaluate i) depression-related immunometabolic dysfunction over time and across illness stages; ii) test whether dysfunction is evident during the disease prodrome; and iii) establish the extent to which peripheral immunometabolic dysfunction relates to brain structure in depression. Figure 1 provides a schematic overview of our study.

**Figure 1.**
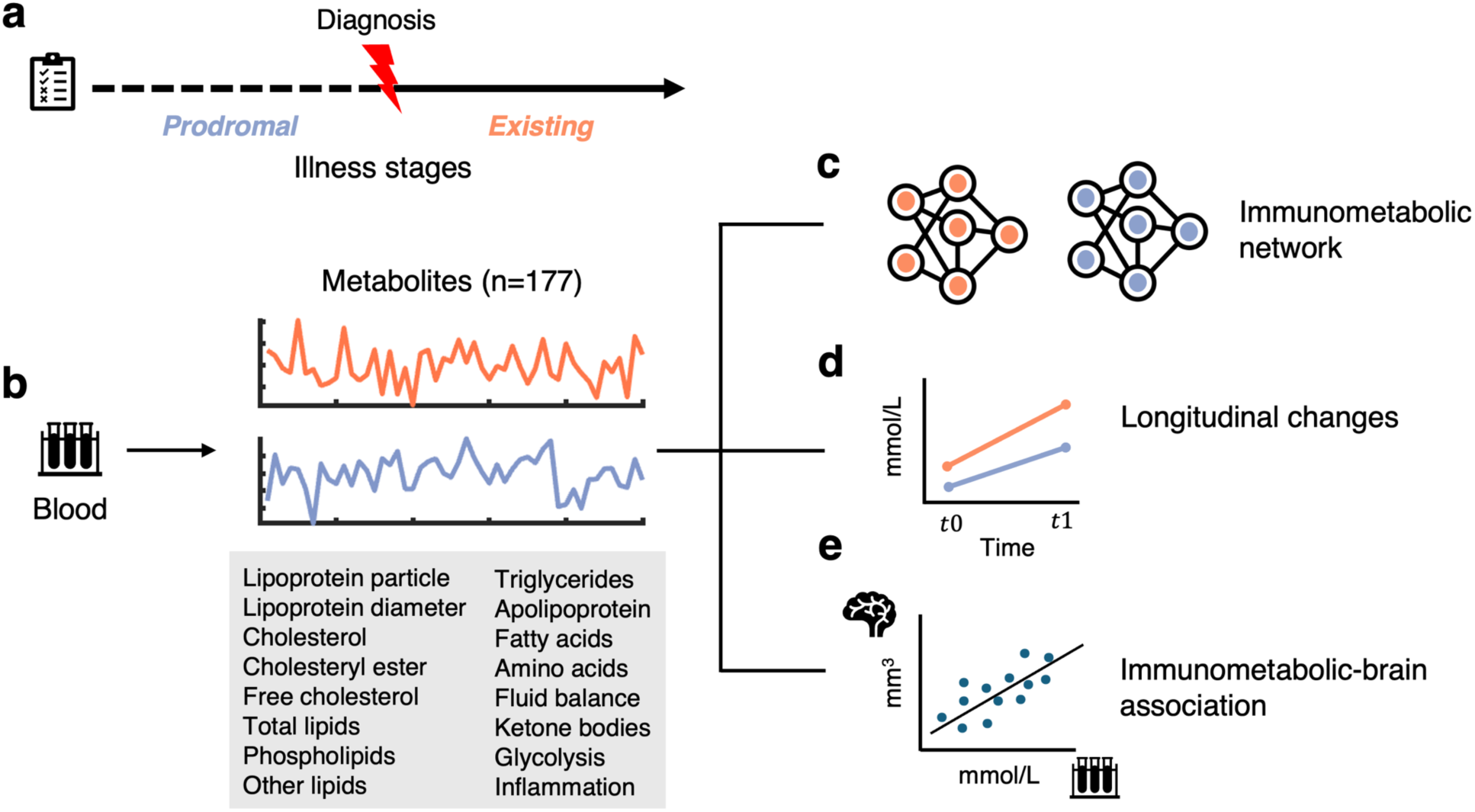
Overview of study design. a), Illness stages at the time of baseline blood sample collection were determined based on health records and self-report. Individuals diagnosed with depression at, or prior to baseline assessment, formed an existing depression group. Individuals diagnosed with depression afterwards defined a prodromal depression group. **b)**, Plasma levels of 177 immunometabolic markers in the existing and prodromal depression groups were compared to healthy individuals. **c),** Schematic showing alterations in immunometabolic networks in depression compared to healthy individuals. The immunometabolic networks were mapped by computing the Pearson correlation coefficient between pairs of metabolites. **d),** Rate of change in the plasma level of each metabolite was computed based on two repeated assessments over a time interval of 2-6 years. **e**), Schematic showing associations between depression-related peripheral immunometabolic signatures and brain gray matter volume.

## Results

We studied a subset of UK Biobank participants with measures of blood-based inflammatory and metabolomic markers. Specifically, 170 peripheral lipid and metabolite biomarkers measured using the Nightingale Health proton nuclear magnetic resonance (NMR) spectroscopy platform were available for 259,367 individuals (age range: 37-71 years, mean 57.1 ± 8.1; 118,708 males (45.8%)) at baseline. Repeat assessment of metabolomic function was available for 15,053 of these same individuals (age range: 43-76 years, mean 62.0 ± 7.4; 7,373 males (49.0%)) during the second visit at 2-6 years follow-up (mean 4.3 ± 0.9 years). Seven blood-based biomarkers derived from standard clinical assays indicating peripheral inflammation were also measured at both baseline and the second visit (Supplementary Table 1). The time interval between consumption of food/drink and blood sample collection (i.e., fasting time) ranged between 0 to 48 hours (mean: 3.8±2.4 hours). Structural brain phenotypes (Supplementary Table 2) were derived from T1-weighted magnetic resonance imaging (MRI) brain scans acquired during the third visit (4-14 years following the initial assessment, mean 9.1 ± 2.0 years) and were available for 27,446 individuals (age range: 45-83 years, mean 64.6 ± 7.7; 13,008 males (47.4%)). A schematic of demographic screening and sample characteristics is provided in Supplementary Figure 1.

Depression diagnostic status was assessed based on multiple clinical records, including primary care, hospital records and death registry, and self-reported questionaries. We identified 45,767 individuals (mean age: 56.1 ± 8.0 years; 16,938 males (37.0%)) who met diagnostic criteria (see Methods) at or prior to the baseline assessment, forming a group that we refer to as the *existing depression* group; 30% with ongoing depressive symptoms at the baseline visit (13,917/45,767) and 9% at both the second (235/2,535) and third (372/4,122) visits.

We defined a *prodromal depression* group that included 24,384 individuals (mean age: 55.5 ± 7.9 years; 8,955 males (36.7%)) who reported depression after the baseline assessment. Among the prodromal depression group, 10,851 individuals had available data showing the date of depression diagnosis. The assessment-to-diagnosis interval ranged between 1 day to 15 years (mean 7.3 ± 3.9 years). We note that the assessment-to-diagnosis interval defined in our study does not necessarily reflect the prodromal phase considered clinically, which is commonly defined as the period between the occurrence of prodromal symptoms (e.g., mild symptoms of anxiety, tension, irritability and somatic) and a depression episode ^46,47^.

Prodromal symptoms may occur before or after the baseline assessment. A healthy comparison group (HC, n=27,636, mean age: 53.8 ± 8.0 years; 13,629 males (49.3%)) included individuals with no existing major medical or mental health conditions at baseline and follow-up visits.

A group of individuals with existing metabolic disorders (including diabetes mellitus, dyslipidemia and obesity) but no concurrent mental health conditions was also curated to provide a positive comparison group (n=59,162, mean age: 59.3 ± 7.6 years; 33,005 males (55.8%)).

### Dysregulated immunometabolism in established and prodromal depression

Peripheral immune and metabolic markers were grouped into 16 categories based on conventional biochemical taxonomy, encompassing lipoprotein particle, lipoprotein diameter, cholesterol, cholesteryl ester, free cholesterol, total lipids, phospholipids, other lipids, triglycerides, apolipoprotein, fatty acids, amino acids, fluid balance, ketone bodies, glycolysis and inflammation. We found markedly different immune and metabolomic profiles in individuals with depression, compared to healthy individuals. Specifically, 155 out of 177 markers showed significantly altered peripheral concentration in the existing depression group, controlling for age, sex, fasting time and cholesterol lowering medication use (absolute value of effect size range: |Cohen’s 𝑑|=0.01 to 0.16, *P*<0.05/177=2.8×10^-4^, Bonferroni correction, Figure 2). We found that depression is primarily associated with i) elevated inflammation, as indicated by increased c-reactive protein (CRP), glycoprotein acetyls (GlycA) and leukocyte count; ii) increased plasma level of VLDL, particularly large to very large particles and their lipid composition; iii) increased plasma level of triglycerides across all lipoprotein subclasses; and iv) decreased plasma level of HDL, particularly large to very large particles, and its lipid composition. Plasma levels of apolipoprotein A1 and the degree of unsaturation of fatty acids were also reduced in depression but to a lesser extent than HDL and VLDL. In addition, we also found that depression is associated with reduced albumin, increased creatinine, and altered levels of amino acids and glycolysis related metabolites. These findings implicate significant depression-related changes in immunometabolism.

**Figure 2.**
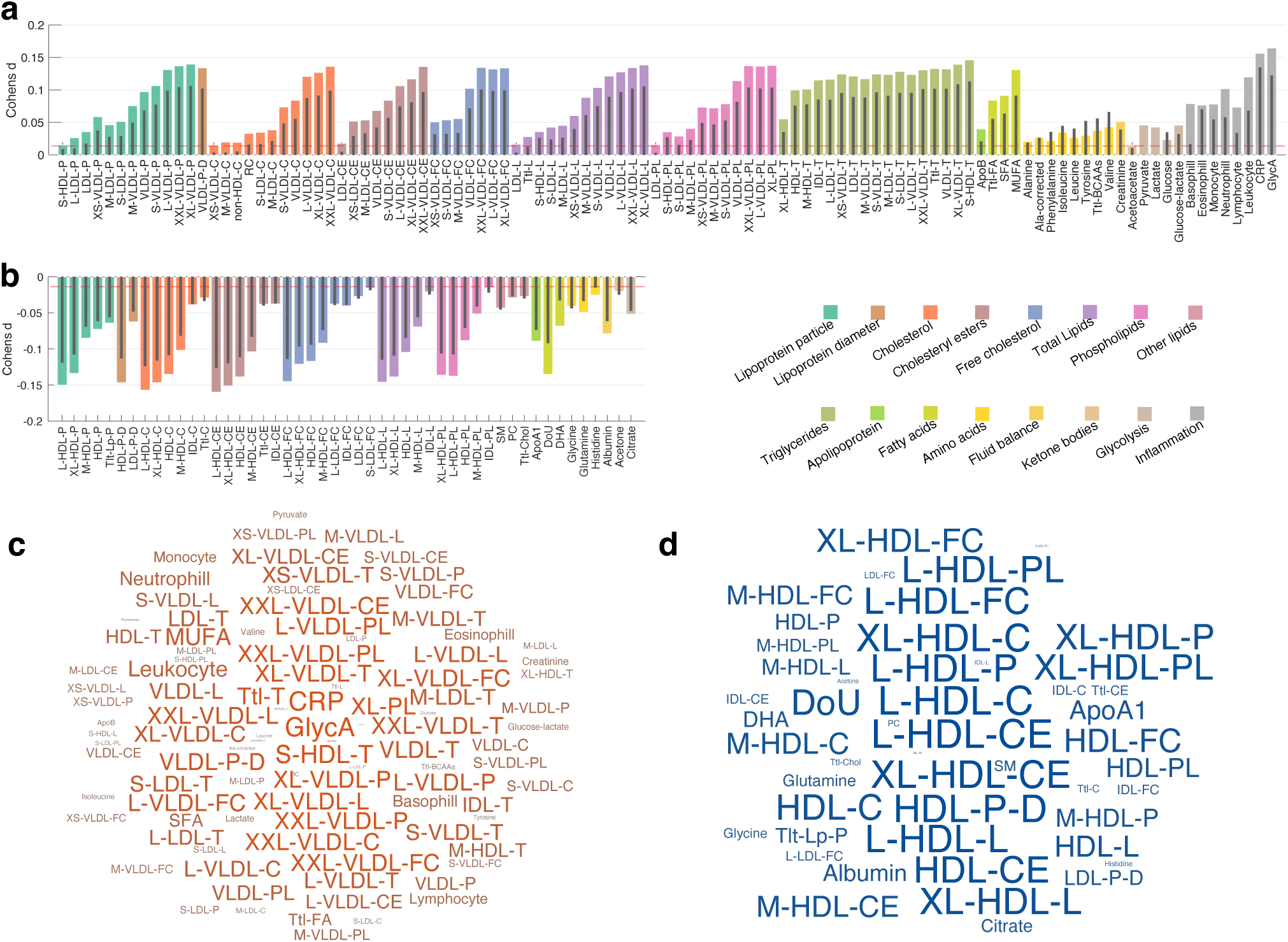
Immunometabolic profiles in depression. Bar plots show effect sizes (Cohen’s 𝑑) for between-group differences in the plasma level of 177 metabolites across 16 metabolite categories. Coloured bars indicate differences between existing depression (n=45,767) and healthy individuals (HC, n=27,636), whereas the inset black lines indicate differences between prodromal depression (n=24,384) and HC. Metabolite categories are shown with uniquely coloured bars. Dashed red line indicates the minimal effect size reaching statistical significance (*P*<0.05/177=2.8×10^-4^, two-tailed, Bonferroni correction). **a)** Metabolites showing higher plasma levels in depression compared to HC. **b)** Metabolites showing lower plasma levels in depression compared to HC. **c,d)**. Word clouds show metabolites that significantly differ between existing depression and HC. The font size is scaled according to the absolute value of the effect size, with red/blue indicating positive/negative Cohen’s 𝑑 values respectively. A full list of abbreviations is provided in Supplementary Table 1.

Importantly, we found that peripheral immune and metabolic dysregulation is evident before the onset of depression (i.e., at the prodromal stage, Figure 2a,b), with 144 out of 177 markers showing significantly altered concentration relative to healthy individuals (|Cohen’s 𝑑|=0.02 to 0.13, *P*<0.05/177=2.8×10^-4^, Bonferroni correction). Although effect sizes in the prodromal group are weaker compared to the existing depression group, the pattern of dysregulation across metabolites is highly consistent between the two groups, as quantified by the Pearson correlation coefficient in Cohen’s d effect sizes across all metabolites (r=0.99, *P*<2.2×10^-308^). A direct comparison between the existing and prodromal group indicated significant but subtle differences in the plasma levels of 132 out of 177 metabolites (|Cohen’s 𝑑|=0.01-0.06, *P*<0.05/177=2.8×10^-4^, Bonferroni correction, Supplementary Figure 2). This suggests that while subtle changes in immunometabolism mark the onset of depression. While our finding does not provide any causal evidence indicating the relationship between immunometabolic dysfunction and depression onset, considerable dysregulation of immune and metabolic markers is observed before illness onset.

We next asked whether the profile of depression-related immunometabolic dysregulation would be comparable to that found in typical metabolic disorders. We found that patterns of inflammatory and metabolomic alterations in the existing depression group show considerable overlap with those found in individuals with metabolic disorders (Supplementary Figure 3). Effect sizes for most metabolites were larger in metabolic disorders compared to depression, but the dysregulation profile is comparable (r=0.97, *P*<2.2×10^-308^). However, several markers including pyruvate, basophil and cholesterol comprised of IDL were slightly less dysregulated in metabolic disorders compared to depression. The plasma levels of 161 metabolites significantly differed between depression and metabolic disorders (|Cohen’s 𝑑|=0.01-0.14, *P*<0.05/177=2.8×10^-4^, Bonferroni correction, Supplementary Figure 4).

A series of supplementary analyses were performed to assess the sensitivity and robustness of our findings, including i) a split-half analysis to assess the reproductivity of the results between randomly split samples (Supplementary Figure 5); ii) further controlling for individual variation in BMI, socioeconomic status, lifestyle, early-life factors and genetic disposition for metabolic function (Supplementary Figures 6 & 7); iii) excluding individuals with established metabolic disorders from the existing and prodromal depression groups (Supplementary Figure 8); iv) excluding individuals who reported regularly taking antidepressants and/or antipsychotics from the existing depression group (Supplementary Figure 9); and v) focusing on depression patients who had received formal clinical diagnosis of major depressive disorder (Supplementary Figure 10). Overall, we found similar patterns of alterations in these inflammatory and metabolomic markers and that most depression-related alterations in metabolites remained significant.

### Systemic metabolomic changes associated with depression

To identify systemic metabolic changes in depression, we mapped metabolomic networks between each pair of metabolites for individuals in the depression and healthy comparison groups. The Pearson correlation coefficient was used to quantify the strength of connection (i.e., coupling) between pairs of metabolites. Age, sex, fasting time and cholesterol lowering medication use were regressed from each metabolite level before correlation analyses. We identified both significantly increased and decreased coupling between metabolites in depression compared to healthy individuals (*P*<0.05/15,576=3.2×10^-6^, Bonferroni correction, Figure 3). Alterations in metabolomic networks were comparable between the existing (Figure 3a) and prodromal (Supplementary Figure 11) depression groups. Specifically, we found that depression is most strongly associated with reduced coupling between glycolysis related metabolites, particularly between glucose and most other metabolite categories (Figure 3c). Increased coupling is also evident in depression, particularly between cholesteryl esters and other lipoprotein and lipid metabolites, and between glucose and alanine amino acid (Figure 3d).

**Figure 3.**
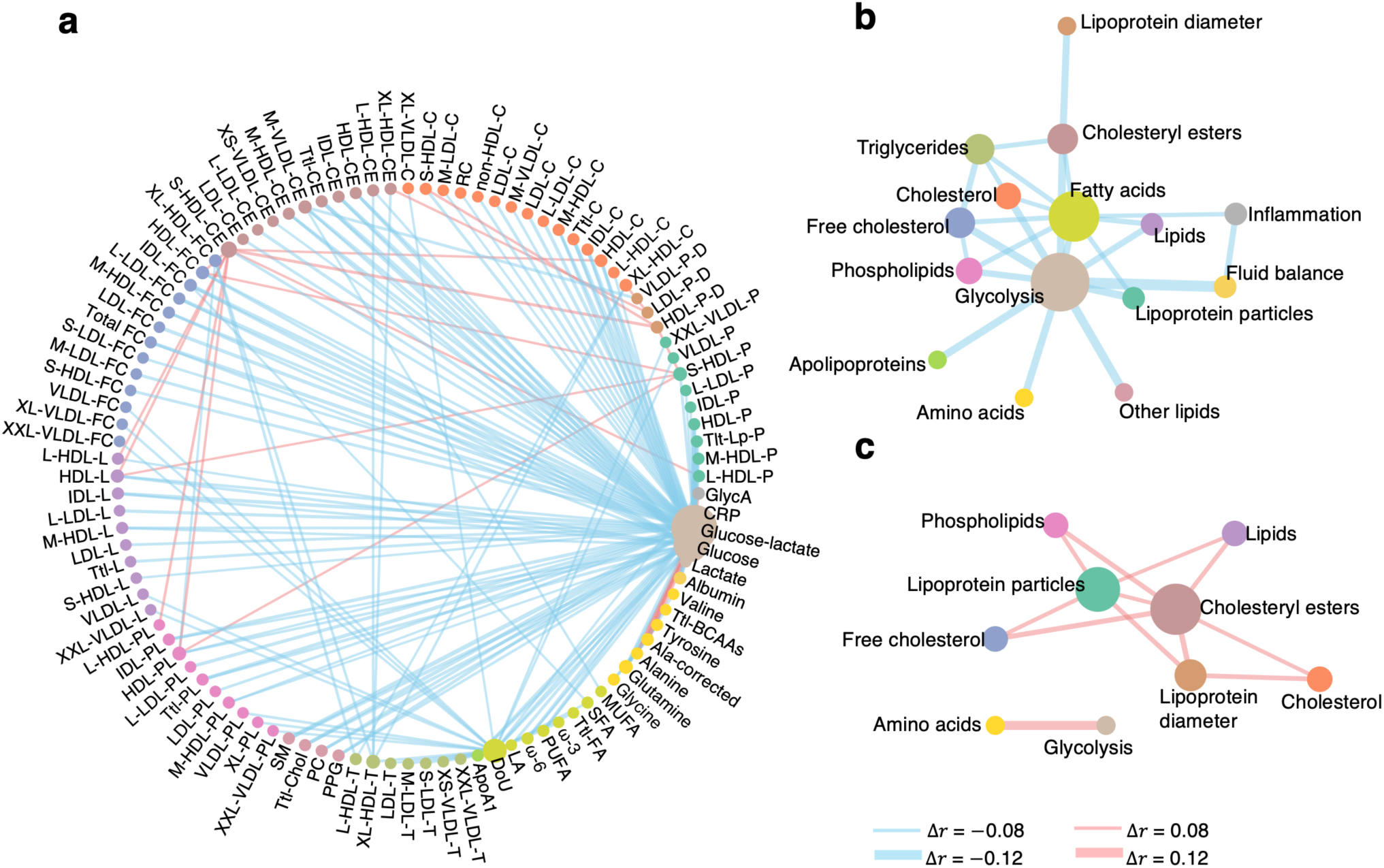
Metabolomic networks in depression. The strength of coupling between pairs of peripheral metabolites was compared between the depression and healthy comparison groups. Networks characterize metabolite pairs with significant between-group differences. **a**) Each network shows the top 1% of connections with significantly altered strength in existing (n=45,767 compared to healthy individuals (n=27,636, *P*<0.05, FDR corrected across 15,576 metabolite pairs). Bootstrapping (n=1000) was used to estimate the distribution of the connectivity strength for each pair of metabolites in each group. **b),** Connections of reduced strength in the same metabolite category were averaged resulting in a summary network showing reduced coupling between metabolite categories in the existing depression group. **c),** Same as panel c but showing increased coupling between metabolite categories. In the graph, blue/red edge represents reduced/increased coupling between metabolites in depression compared to healthy individuals. Node size is modulated by the degree of nodal strength. Nodes of metabolites of the same category were colored the same. Edge thickness is modulated by the altered connectivity strength.

### Longitudinal immunometabolic profiles in depression

We next investigated the extent to which peripheral immunometabolic profiles in depression change as a function of illness progression. We first computed the rate of change per year in the plasma level of each metabolite between baseline and follow-up visits (time interval: 2-6 years) for each individual. The rate of change was estimated as the Δ = *(Xt1* − *Xt0)/T*, where *Xt1* and *Xt0* are the plasma levels of a given metabolite at the follow-up and baseline visits respectively, and *T* is the number of years between the two time points. A larger positive/negative rate implies a fast increase/decrease of metabolites. Age, fasting time, cholesterol lowering medication use and sex were regressed from each metabolite at each study visit respectively before computing the rate of change. One-way analysis of covariance was then used to test whether the rate of change in each metabolite differs between the existing and prodromal depression groups and healthy individuals, controlling for age (average age across two time points) and sex. We found significant between-group differences in the rate of change for glucose (*F*=9.3, *P*=4.9×10^-5^), lactate (*F*=37.9, *P*=4.2×10^-17^), pyruvate (*F*=17.3, *P*=3.1×10^-8^) and basophil count (*F*=9.4, *P*=8.8×10^-5^). The false discovery rate (FDR) was controlled at 0.05 across 177 metabolites using the Benjamini-Hochberg procedure ^48^. Post-hoc analyses using two-sample t-tests revealed a faster increase of glucose (*t*=4.1, *P*=3.4×10^-5^), and a faster decrease of lactate (*t*=8.4, *P*=5.3×10^-17^), pyruvate (*t*=5.7, *P*=9.9×10^-9^) and basophil count (*t*=4.2, *P*=3.3×10^-5^)) in existing compared to prodromal depression. However, only the rate of change in lactate significantly differed between depression and healthy individuals (existing depression vs HC: *t*=4.7, *P*=3.1×10^-6^, Figure 4a). This suggests that the longitudinal rate of change in these markers may be a sensitive indicator of depression progression. However, the relationship between disease progression and the change in these metabolites over time may be non-linear, given the absence of a consistent pattern distinguishing individuals with depression from healthy controls.

**Figure 4.**
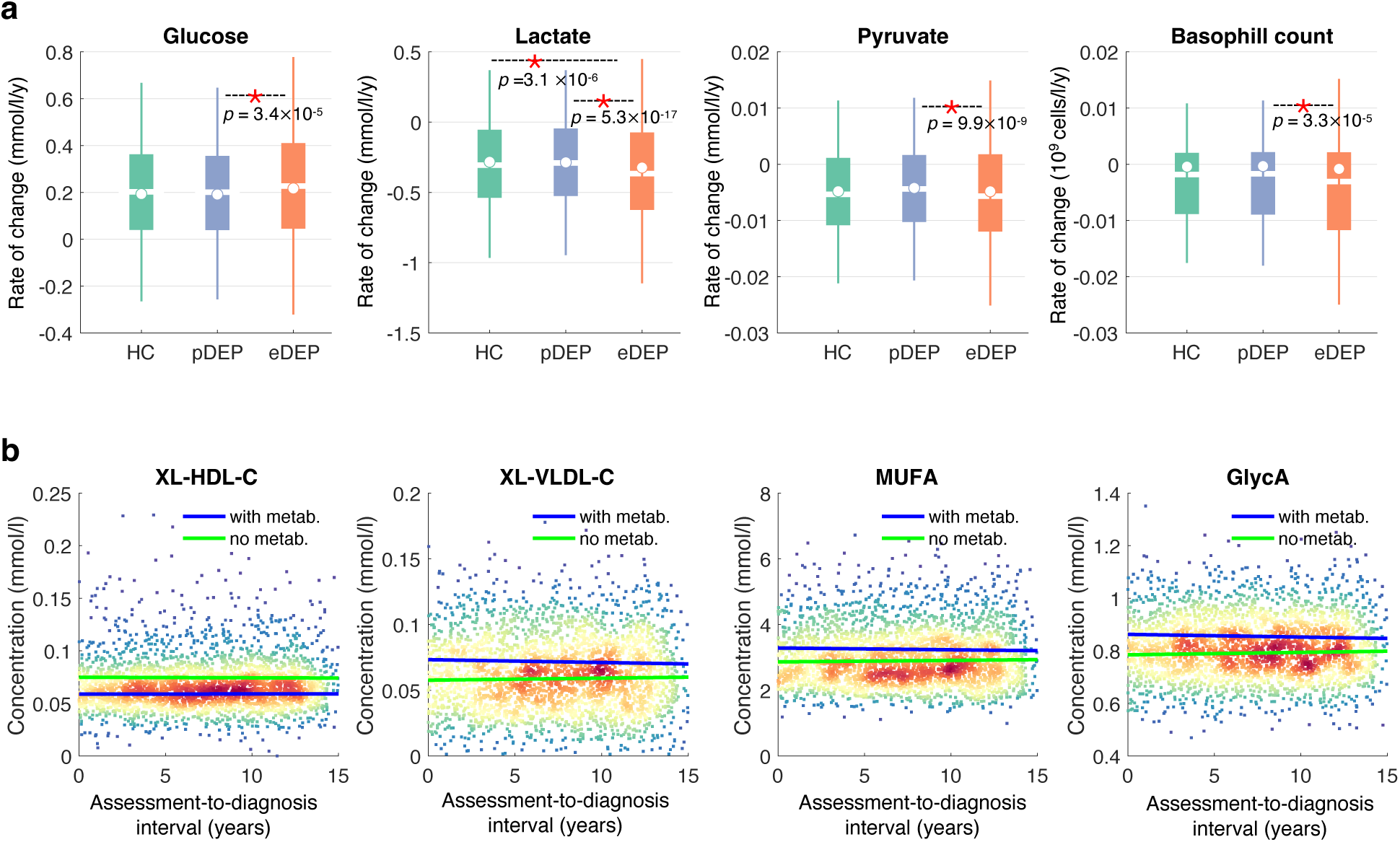
Metabolomics over time in depression. a), Metabolites that differed significantly in the mean rate of change per year between existing depression (eDEP, n=2,535), prodromal depression (pDEP, n=3,045) and healthy comparison individuals (HC, n=1,208). Fasting time and cholesterol medication use were first regressed from each metabolite at each time point. Analysis of variance was then used to test for between-group differences for each metabolite, controlling for sex and average age across the two time points. The asterisk indicates significantly different mean rate of change between two groups using a two-sample t-test. The bottom and top edges of the boxes indicate the 25th and 75th percentiles of the distribution, respectively. The central line indicates the median and the dot indicates the mean. The whiskers extend to the most extreme data points that are not considered outliers (1.5-times the interquartile range). **b),** Variation in plasma levels of representative metabolites over the assessment-to-diagnosis interval. Lines of best fit show associations between assessment-to-diagnosis interval and metabolite levels in individuals with (blue) and without (green) comorbid metabolic disorders (metab.). XL-HDL-C, very large high-density lipoprotein cholesterol; XL-VLDL-C, very large very-low-density lipoprotein cholesterol; MUFA, monounsaturated fatty acids; GlycA, glycoprotein acetyls.

We then assessed the extent to which peripheral immunometabolic profiles in prodromal depression vary as a function of the assessment-to-diagnosis interval. However, we did not find any significant associations between plasma levels of metabolites and the time interval between baseline blood sample collection and depression diagnosis, controlling for age, sex, fasting time, cholesterol lowering medication use (*P*>0.05, FDR correction across 177 metabolites). The metabolic disorders × assessment-to-diagnosis interval interaction did not show significant effects on the plasma levels of metabolites (*P*>0.05, FDR correction across 177 metabolites), indicating that the association between metabolites and the assessment-to-diagnosis interval did not differ between individuals with and without comorbid metabolic disorders. These findings suggest that immunometabolic dysregulation is evident years before the onset of depression and remains relatively stable over time. Figure 4b shows the stable trajectories of several representative immunometabolic markers over the assessment-to-diagnosis interval.

### Immunometabolic-brain associations in depression

Finally, we investigated whether depression-related immunometabolic dysregulation associates with brain structure in the disorder. To this end, we first derived three summary measures of metabolites using principal component analysis in the entire sample of individuals with available metabolites data (Figure 5a). The first three principal components explained 36.9%, 23.3% and 13.5% of the total variance respectively. We assessed the main effects of peripheral metabolites and depression diagnosis as well as the diagnosis × metabolites interaction effect on brain gray matter volume for each brain region and for each of the three principal components of metabolites, controlling for age at blood sample collection, age at brain scan, sex, fasting time and cholesterol lowering medication use. For the main effect of depression diagnosis (n=4,122) relative to healthy individuals (n=2,558), we only found a significant association between depression and reduced volume of ventral diencephalon (Cohen’s 𝑑=-0.05, *P*=1.2×10^-4^, FDR corrected across 42 brain regions × 3 metabolite components = 126 tests). The effect sizes of regions that showed nominal significance (*P*<0.05), including the inferior parietal lobule, medial orbitofrontal cortex, banks of the superior temporal sulcus, middle temporal gyrus, thalamus, pallidum and the cerebellum are shown in Supplementary Figure 12. We did not observe significant main effects of metabolites on brain gray matter volume. However, a significant interaction effect was evident for paracentral (Cohen’s 𝑑=-0.044, *P*=3.2×10^-4^) and superior parietal (Cohen’s 𝑑=-0.042, *P*=6.9×10^-4^, FDR corrected across 42 brain regions × 3 metabolite components = 126 tests) lobules in the association with the second metabolite principle component (PC2) (Figure 5b), which primarily represents low circulating albumin and high CRP. Specifically, we found that each one standard deviation increase in PC2 score is associated with 40.6mm^3^ and 124.2 mm^3^ reduction in the paracentral and superior parietal lobule gray matter volume respectively in the existing depression group (Figure 5c). There was no significant association between PC2 score and the regional gray matter volume in healthy individuals. Controlling for comorbid metabolic disorders in depression did not change the significance of the results. In a post-hoc analysis, the main effect of diagnosis on ventral diencephalon volume (Cohen’s 𝑑=-0.06; p=5.3×10^-6^) and the interaction effects of diagnosis × metabolites on the gray matter volume of paracentral (Cohen’s 𝑑=-0.037; p=0.002) and superior parietal (Cohen’s 𝑑=-0.034; p=0.005) lobules remained significant after further controlling for individual variation in intracranial volume. We repeated the analysis using cortical thickness measures, however; we did not find a significant diagnosis × metabolites interaction effect on cortical thickness (*P*>0.05, FDR correction across 33 cortical brain regions × 3 metabolite components = 99 tests).

**Figure 5.**
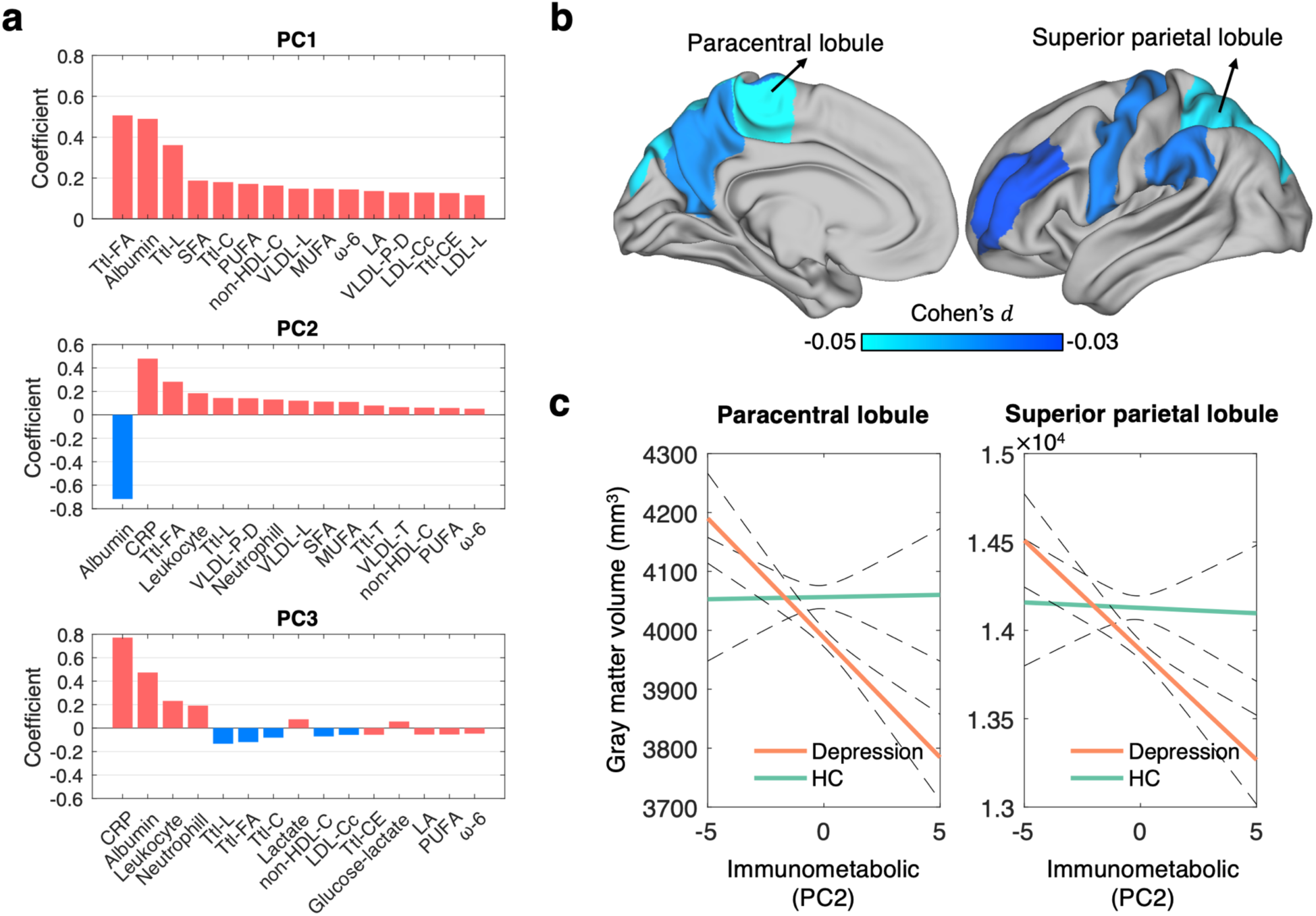
Metabolomic-brain associations in depression. a), Bar plots show the loading (coefficients) of 15 top-ranked markers on the each of the first three principal components of the peripheral metabolites. **b)**, Effect sizes (Cohen’s d) of significant depression diagnosis × metabolites interaction effect on paracentral (Cohen’s 𝑑=-0.044, *P*=3.2×10^-4^, FDR corrected) and superior parietal (Cohen’s 𝑑=-0.042, *P*=6.9×10^-4^, FDR corrected) lobules. Cortical regions that showed nominally significant interaction effects are also rendered on the cortical surface (Desikan-Killiany atlas) for visualization. These include precentral (Cohen’s 𝑑=-0.032, *P*=0.0009), precuneus (Cohen’s 𝑑=-0.034, *P*=0.006), rostral middle frontal (Cohen’s 𝑑=-0.026, *P*=0.03), supramarginal (Cohen’s 𝑑=-0.031, *P*=0.01). In addition, amygdala (Cohen’s 𝑑=-0.024, *P*=0.04) and the total gray matter volume (Cohen’s 𝑑=-0.025, *P*=0.04) were also nominally significant (not shown). **c)**, Lines of best fit show associations between metabolites (PC2) and gray matter volume for paracentral and superior parietal lobules in established depression (orange) and healthy individuals (green) separately.

## Discussion

Using a large-scale prospective population cohort, we evaluated peripheral immunometabolic dysfunction in depression over time and across illness stages. Integrating brain imaging and comprehensive immunometabolic data, our study is the largest of its kind in depression. We revealed relatively persistent patterns of immunometabolic dysfunction before and after the onset of depression and over the illness course, irrespective of the presence of comorbid metabolic disorders and the use of psychotropic medications. We identified systemic changes in metabolomics and a prospective association between peripheral immunometabolic dysfunction and brain structure in depression. Our work establishes both population-level and individual-level immunometabolic dysregulation profiles associated with depression. It highlights the importance of managing chronic low-grade inflammation and altered lipid and glucose metabolism in the disorder.

Consistent with previous studies ^11–13^, we report significantly altered peripheral lipid profiles in individuals with depression, evidenced by increased circulating VLDL and triglycerides and decreased HDL. We found that among lipoprotein subclasses, very large and large sized particles tend to have a stronger association with depression compared to smaller particles. For example, the concentration of small HDL and its cholesterol composition were not significantly associated with depression, which may be because a portion of these are nascent HDL and represent newly synthesized particles. However, the remainder may arise from dysfunctional remodelling in response to pathological processes, including inflammation ^49^. In contrast, larger sized HDL plays a central role in reverse cholesterol transport by carrying cholesterol back to the liver for disposal ^50^. Indeed, whereas large HDL is often associated with reduced risk of cardiovascular diseases, the association with small HDL remains inconclusive ^51^. Similarly, we found a several-fold weaker association between depression and small sized VLDL compared to larger VLDL. The small sized VLDL, which represents the final stage of triglyceride-rich lipoprotein metabolism, may be less involved in the pathology of depression. Although routine clinical assays do not assess lipoprotein subclasses, our finding suggests the potential advantage of using these biomarkers in assessing metabolic disorder risk in depression, particularly at early stages of illness given their superior sensitivity. Moreover, compared to BMI, a proxy of general metabolic health, assessing these immunometabolic markers may enable identification of specific metabolic pathways that are affected in depression. Despite the small-to-moderate effects sizes, our findings are robust and highly reproducible in random split samples. The coordinated action of multiple metabolites may result in a greater cumulative impact on disease risk and symptomatology, extending beyond the effect of any individual metabolite. Further work investigating the clinical relevance of the combined effects are required.

We found that the altered peripheral inflammatory and lipid profiles were evident before the onset of depression (7-year prior on average). Whereas existing studies focus on individuals with established depression ^11–14^, our study provides new evidence of immunometabolic dysregulation well before illness onset. Although the causal relationship between immunometabolic dysfunction and depression onset remains to be established, our findings suggest that metabolomic dysfunction may confer risk of subsequent depression. However, it is noteworthy that the UK Biobank comprises middle aged and older individuals, who are more prone to age-related immunometabolic dysfunction compared to younger adults. Further work including a cohort of young adults is needed to provide a complete characterization of the interrelationship between immunometabolic dysfunction and depression across the adult lifespan.

Interestingly, we found that preexisting metabolic dysfunction in prodromal depression remained relatively persistent over the illness course and was not markedly exacerbated by illness onset. In fact, differences in inflammatory, lipid and lipoprotein profiles between prodromal and existing depression were relatively subtle. Whereas the immunometabolic markers were generally less affected in prodromal depression compared to the existing depression group, the alteration pattern across the entire immunometabolic panel was highly consistent between the two, and thus suggests system-level immunometabolic dysregulation. Moreover, we did not observe significant associations between the plasma levels of immunometabolic markers and the assessment-to-diagnosis interval. This suggests that alterations in immunometabolic markers may have been established earlier and are less sensitive to the current depression state. However, when assessing these markers longitudinally, several glycolytic markers including glucose, lactate and pyruvate as well as basophil count showed faster rate of change in existing depression compared to the prodromal group. This suggests that glycolysis and innate and adaptive immune dysfunction in depression are not only associated with depression pathology ^52^ but are also associated with the progression of the illness, and possibility depression-related accelerated aging ^53^. A longitudinal study design that follows depression patients from prodromal to established depression will be needed to better characterize the relationship between metabolites changes and the clinical trajectory of depression symptomology over time.

Moreover, we found widespread alterations in the strength of coupling between pairs of metabolites in depression compared to healthy individuals. Altered glycolysis coupling was most strongly implicated in the network analysis. Although the underlying mechanisms explaining these network-level alterations remains to be addressed, our findings suggest systemic depression-related changes in metabolites, and that glycolysis is central to these systemic changes. Glycolysis is a critical pathway in cellular energy production and decoupling of glycolysis from other metabolic pathways may reflect disrupted energy metabolism. Under inflammatory conditions, immune cells are known to shift energy metabolism away from oxidative phosphorylation and towards glycolysis, as process known as the Warburg effect ^54^. It is also consistent with the notion that peripheral metabolic stressors (e.g., cholesterol crystals, oxidised phospholipids) could induce glycolytic reprogramming in activated myeloid cells in association with proinflammatory responses ^55,56^, contributing to systemic inflammation. Nevertheless, further work is needed to determine whether interventions targeted at normalizing glycolysis regulation would facilitate downstream improvement of systemic immunometabolic dysfunction and symptom alleviation in depression.

Finally, consistent with the existing literature ^27–30^, we found that depression is associated with reduced gray matter volume across multiple brain regions, particularly those in frontal, temporal and subcortical areas; however, the effect sizes found here are weaker compared to those reported previously and some of them are only nominally significant. The weaker effect sizes may be explained by the symptom remission status of depression individuals included in our study, as most of them (>90%) had no or very mild depressive symptoms at the time of brain scans (PHQ-2 < 3). Importantly, we found that immunometabolic dysfunction relates to smaller brain gray matter in depression. The peripheral immunometabolic signature that contributes to reduced brain gray matter volume was primarily driven by low plasma levels of albumin and high levels of CRP. Whereas albumin has anti-inflammatory properties, higher levels of CRP indicate elevated systemic inflammation. Our finding is thus consistent with the inflammation hypothesis of depression ^57^, whereby peripheral inflammation may induce neuroinflammation and subsequent brain changes and depressive symptoms. The absence of this immunometabolic-brain association in healthy individuals further supports the potential pathological and negative impact of peripheral immunometabolic disruption on the brain. However, the two brain regions (i.e., paracentral and superior parietal lobules) that most strongly associate with peripheral immunometabolic dysfunction in depression did not overlap with the brain regions that show differences in volume between depression and healthy individuals (e.g., ventral diencephalon, medial orbitofrontal cortex, superior temporal gyrus). Whereas the paracentral lobule, as well as medial orbitofrontal cortex and superior temporal gyrus are parts of the default mode network, the superior parietal lobule is part of the fronto-parietal network, and the disturbances of both circuits have been widely implicated in depression ^58^. Further work integrating peripheral and central inflammatory markers as well as brain connectivity is needed to investigate these effects.

This study has several limitations. First, overnight fasting was not required prior to blood sample collection in the UK Biobank. Peripheral lipid and glucose profiles could thus be confounded by food intake. Although individual variation in fasting time was controlled in all statistical analyses, where applicable, further work validating our findings in an independent cohort with consistent overnight fasting is needed. Second, despite the rich set of lipoprotein and lipid biomarkers, the availability of peripheral inflammatory markers is relatively limited. Future work including peripheral cytokines would provide greater insight into the complex interplay between immunometabolic dysregulation and neuropathology associated with depression. Third, detailed clinical assessment and treatment response information is not available for the UK Biobank cohort. We were thus unable to assess the extent to which peripheral immunometabolic dysfunction influenced the clinical trajectory of depression, or if there is a dose-dependent relationship between depressive symptoms severity and immunometabolic dysfunction. Fourth, due to the sequential and non-randomized participant assessment, we were unable to assess the extent to which metabolic function may be influenced by depression-related brain structural and functional changes. Further work is also needed to test if individual variation in brain structure observed at follow-up is consistent with alterations in brain structure assessed longitudinally, consequent to alterations in metabolites. Immunometabolic dysfunction could also manifest in other psychiatric disorders and not depression specifically. The extent to which immunometabolic dysfunction is a transdiagnostic marker remains to be assessed. Finally, the UK Biobank only comprises middle-to-old-aged participants. The extent of immunometabolic dysfunction and its manifestation on brain structure in young adults with depression remains to be addressed.

In conclusion, our work reveals persistent and systemic changes in peripheral immunometabolic profiles in depression over time and across illness stages. Our findings suggest that disturbances in peripheral immunometabolic regulation may confer the risk of depression development and influence brain structure underpinning the neuropathology of depression. Early detection and intervention focussed on normalizing immunometabolic dysfunction may mitigate the risk of depression and improve depression prognosis.

## Methods

### Dataset

Blood-derived metabolomics and inflammatory biomarkers and brain imaging data analysed in this study were sourced from the UK Biobank. The UK Biobank is a large-scale biomedical database and research resource containing genetic, lifestyle and health information from approximately 500,000 participants^44,45^. The UK Biobank has approval from the North West Multi-centre Research Ethics Committee (MREC) to obtain and disseminate data and samples from the participants (http://www.ukbiobank.ac.uk/ethics/). Written informed consent was obtained from all participants.

### Inflammatory and metabolomics biomarkers

A total of 275,256 individuals (age range: 37-73 years, 126,753 males) had NMR-based metabolomics data ^12^ based on plasma samples collected at baseline (2006-2012). Among these individuals, 18,857 individuals (age range: 43-79 years, 9,259 males) had a repeat assessment 2-6 years later (2012-2013). The NMR technique provides a quantification of plasma concentration of 170 metabolites and 81 derived ratio indices. In this study, we considered the 170 concentration measures. In addition, seven peripheral inflammatory biomarkers measured using standard clinical hematology and biochemistry assays were also considered, resulting in 177 biomarkers in our present study (Supplementary Table 1).

Details of UK Biobank blood sample collection, processing and storage protocol are provided elsewhere ^59^. Following a previous study ^11^, for each biomarker, a value that resides more than 5 standard deviations away from the median was considered an outlier. Individuals with any outlier values were excluded, resulting in a final sample of 259,367 individuals (age range: 37-71 years, mean 57.1 ± 8.1, 118,708 males) at baseline and 15,053 individuals (age range: 43-76 years, mean 62.0 ± 7.4, 7,373 males) at follow-up.

Individuals who had follow-up metabolomics data were slightly older at baseline compared to those who did not have follow-up metabolomics data (mean age: 57.7 ± 7.4 years vs 57.0 ± 8.1 years; Cohen’s d=0.01, *P*=3.2×10^-22^). There was also a slightly higher proportion of male participants among individuals who had follow-up metabolomics data (7,373/15,053=0.48) compared to those who did not (111,343/244,333=0.46, Chi-square=66.4, *P*<0.00001).

### MRI-derived brain phenotypes

Multimodal brain imaging was acquired during the third visit (2014-2020) ^45^ and were available for 27,446 individuals (age range: 45-83 years, mean 64.6 ± 7.7, 13,008 males). Regional cortical (Desikan-Killiany atlas ^60^) and subcortical gray matter volume derived from T1-weighted MRI were sourced from the UK Biobank. Details of the image processing pipeline, artefact removal, cross-modality and cross-individual image alignment, quality control and phenotype estimation are described elsewhere ^61^ and are available in the UK Biobank brain imaging documentation (https://biobank.ctsu.ox.ac.uk/showcase/showcase/docs/brain_mri.pdf). Regional measures were averaged across the left and right hemispheres, resulting in 42 gray matter regional measures (Supplementary Table 2). As we did not have a strong hypothesis on laterality, averaging regional measures across hemispheres improved the signal-to-noise ratio of MRI phenotypes and reduced the number of multiple comparisons.

### Health records and clinical information

Medical information and clinical diagnosis were obtained through self-report (UK Biobank field ID: 20002) and data linkage to health care records from the UK National Health Services (field IDs: 41270; 41271; 42040), which included International Classification of Diseases and Related Health Problems (ICD-10) coded major depressive disorder, single episode (F32) and major depressive disorder, recurrent (F33). A probable depression status was considered if an individual reported yes to questions of ever depressed or unenthusiastic/disinterested (field ID 4595 or 4631) for at least a week (field ID 4609 or 5375) and had been seen by a general practitioner or psychiatrist for the above (filed ID 2090 or 2100). At each visit, an individual’s current depression status was assessed by a 2-item depression subscale (frequency of depressed mood in last 2 weeks, field ID 2050; frequency of unenthusiasm/disinterest in last 2 weeks, field ID 2060) of a 4-item Patient Health Questionnaire (PHQ-4) ^62^. We re-coded the answer to each question to 0-not at all, 1-several days, 2-more than half the days and 3-nearly every day, to be consistent with the original scale. A sum score of 3 or more across the two items is considered depression ^63^. Given the heterogeneity in coding practices and known underdiagnosis of depression in community populations ^64^, an absence of an ICD-10 depression diagnosis does not necessarily exclude a diagnosis of depression Including probable depression allows increases the likelihood of detecting individuals who may have undiagnosed clinical depression at the time of assessment.

A group of individuals with metabolic disorders including diabetes mellitus (E10-E14), dyslipidemia (E78) or obesity was defined based on clinical records of diagnosis and body mass index>30.

Medication information of cholesterol lowering medications, antidepressants and antipsychotics were obtained through self-report (field ID 20003). The proportions of individuals who were taking cholesterol lowering medications are 8,256/45,767=0.18, 3,423/24,384=0.14, and 25,153/59,162=0.42 in the existing depression, prodromal depression and metabolic disorders groups, respectively. The proportions of individuals who were taking antidepressants and/or antipsychotics were 0.29 (13,085/45,767) and 0.02 (767/45,767) respectively in the existing depression group. The list of drugs included in each category is provided in Supplementary Table 3.

### Additional covariates

Supplementary analyses assessed BMI, and 14 potential confounding variables including socioeconomic status, lifestyle, early-life factors and genetic disposition for metabolic dysfunction.

- BMI (Field ID 21001): Body mass index defined as the body weight divided by the square of the body height in units of kg/m^2^.
- Social economic status was indexed by Townsend deprivation index (Field ID 22189).
- Responses to current tobacco smoking (Field ID 1329) was recoded to 0=No; 1=only occasionally; 2=Yes, on most or all days, so that higher score denoted higher frequency of current smoking.
- An average weekly alcohol consumption (in UK standard units) was computed by combing information on each person’s questionnaire response on weekly and monthly intake of a variety of beverage type, including red wine, white wine/champagne, spirits and fortified wine (Field IDs 1568, 1578, 1588, 1598, 1608, 4407, 4418, 4429, 4440, 4451). Specifically, weekly alcohol intake data were collected from individuals who indicated that they drink more often than once or twice a week, whereas monthly alcohol intake was collected from individuals who drink alcohol one to three times a month or on special occasions. The alcohol consumption of individuals who indicated that they never drink (Field ID: 20117) was set to zero.
- Usual walking pace (Field ID 924): 1=slow pace; 2=steady average pace; 3=brisk pace.
- Summed MET minutes per week (Field ID 22040): Total Metabolic Equivalent Task (MET) minutes per week for all activity including walking, moderate and vigorous activity.
- Breastfed as a baby (Field ID 1677): 1=Yes; 0=No.
- Comparative body size at age 10 (Field ID 1687): 1=Thinner; 2=Plumper; 3=About average
- Comparative height size at age 10 (Field ID 1697): 1=Shorter; 2=Tailer; 3=About average
- Maternal smoking around birth (Field ID 1787): 1=Yes; 0=No.
- Standard polygenetic risk scores of BMI (Field ID 26212), HDL-cholesterol (26242), LDL-cholesterol (26250) and type 2 diabetes (26285).

### Mapping metabolomic networks

The Pearson correlation coefficient was used to quantify the strength of connectivity between pairs of metabolites, controlling for age, sex, fasting time and cholesterol lowering medication use, in the existing depression, prodromal depression and healthy comparison group separately. To assess between-group differences in connectivity strength, we first estimated the distribution of the correlation coefficient for each metabolite pair in each group separately using bootstrapping (n=1,000). In each bootstrapped sample, connectivity strength was re-computed for each metabolite pair, controlling for the correspondingly resampled covariates. An Anderson-Darling test ^65^ was used to test the null hypothesis that the connectivity strength between metabolites in the bootstrapped sample is from a population with a normal distribution. We found that data normality was met for all connection pairs (*P*≥0.05, FDR corrected across 15,576 pairs of metabolites) in all three groups. A two-sample t-test was then used to assess whether the connectivity strength significantly differed between healthy individuals and the two depression groups respectively. The FDR was controlled at 0.05 across 15,576 pairs of metabolites.

A permutation testing (n=10,000) was also used to determine the significance of between-group differences for each metabolite pair. The p-value was given by the proportion of permutations with the t-statistic that exceeded or equalled the t-statistic in the unpermuted data. Permutation testing is non-parametric and does not require any assumptions on the distribution of samples and minimal assumptions about homoscedasticity. The results across the two statistical testing approaches were consistent.

## Data Availability

Data were obtained from the UK Biobank. Researchers can register to access all data used in this study via the UK Biobank Access Management System (https://bbams.ndph.ox.ac.uk/ams/).

## Acknowledgments

This research has been conducted using data from UK Biobank (https://www.ukbiobank.ac.uk/), a major biomedical database. We that the UK Biobank for making the data available, and to all study participants, who generously donated their time to make this resource possible. YET is supported by a National Health and Medical Research Council Investigator Grant (APP2026413). AZ is supported by a Future Fellowship from the Australian Research Council (FT220100091).

## Author Contributions

YET conceived the idea, designed the study, compiled the data, performed the analyses, prepared the visualizations and drafted the manuscript. AZ, CG, MAD, RC and VC provided critical conceptual and technical input. All authors provided critical feedback and editing of the final manuscript.

## Code availability

Custom Matlab code (R2022b, Natick, Massachusetts: The MathWorks Inc.) was used to perform statistical analyses and data visualization. Brain renderings were created using the Connectome Workbench software (https://www.humanconnectome.org/software/connectome-workbench).

