## Supplementary Information for "Immunometabolic dysregulation in depression predates illness onset and associates with lower brain gray matter volume"

**Table of Contents**

Figure S1. Schematic of sample characteristics.

Figure S2. Comparison of immunometabolic markers between existing and prodromal depression groups.

Figure S3. Immunometabolic profile in metabolic disorders.

Figure S4. Comparison of immunometabolic markers between metabolic disorders and existing depression groups.

Figure S5. Reproducible immunometabolic profiles in depression in split-half samples.

Figure S6. Altered immunometabolic profiles in depression controlling for individual variation in socioeconomic status, lifestyle, early-life factors and genetic disposition for metabolic function.

Figure S7. Altered immunometabolic profiles in depression controlling for individual variation in BMI, socioeconomic status, lifestyle, early-life factors and genetic disposition for metabolic function.

Figure S8. Immunometabolic profiles in depression without established metabolic disorders.

Figure S9. Immunometabolic profiles in psychotropic-medication naïve depression.

Figure S10. Altered immunometabolic profiles in clinically diagnosed depression.

Figure S11. Metabolomic networks in prodromal depression

Figure S12. Brain gray matter volume reduction associated with depression.

Supplementary Tables 1-3 are provided in an excel format.

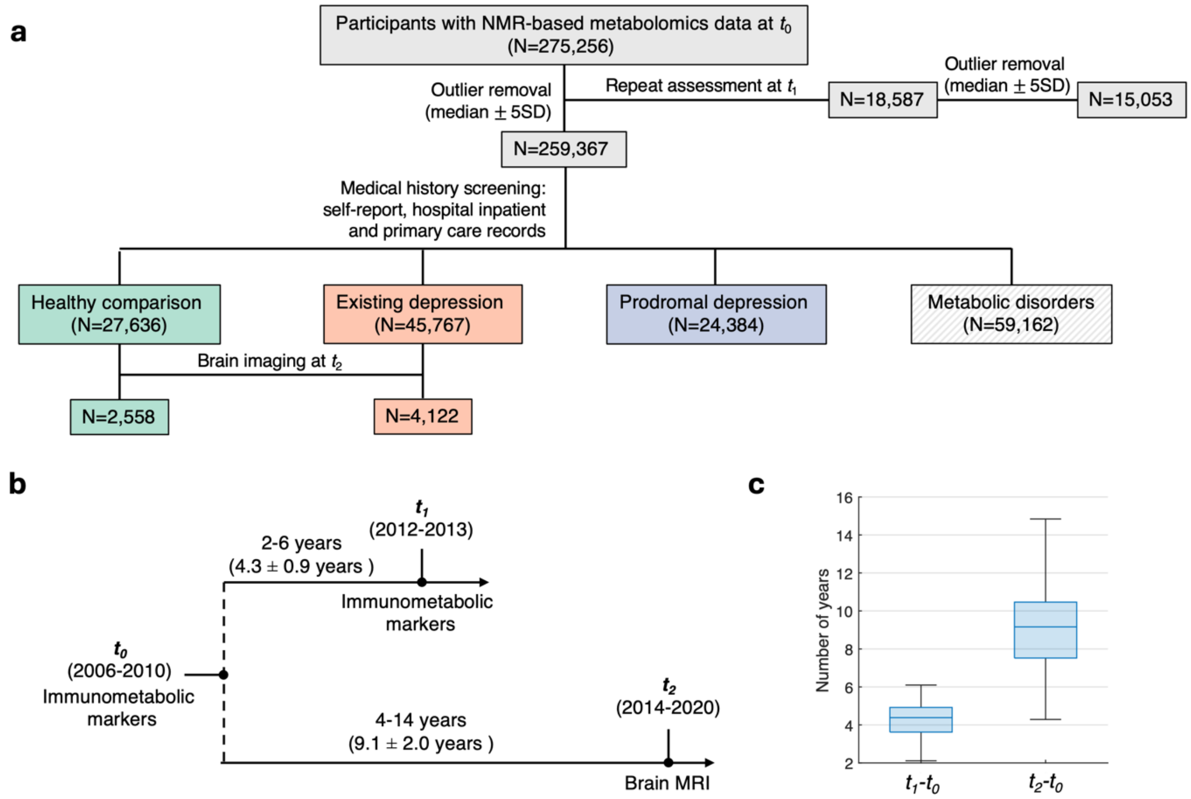


**Figure S1**. Schematic of sample characteristics. **a)** Inclusion and exclusion criteria for each analysis group at each available study visit; **b)** time of assessment for blood-based immunometabolic markers and brain MRI; **c)** individual variation in the time interval between baseline ($t_0$) and repeated assessment for blood markers at the second study visit ($t_1$) and brain MRI at the third study visit ($t_2$). NMR, nuclear magnetic resonance spectroscopy; SD, standard deviation. MRI, magnetic resonance imaging.

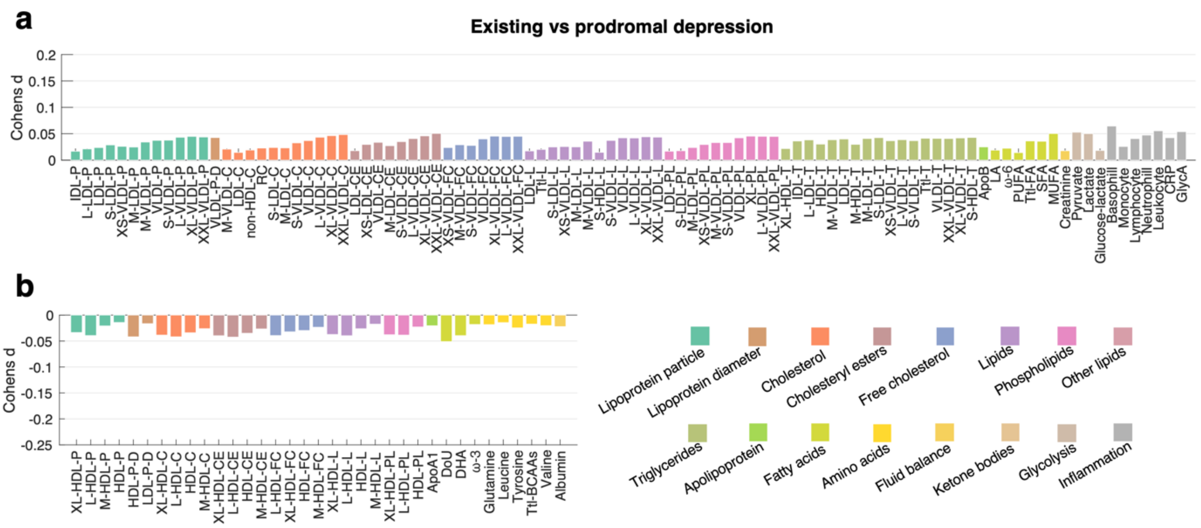


**Figure S2. Comparison of immunometabolic markers between existing and prodromal depression groups.** Bar plots show effect sizes (Cohen's $d$) of metabolites showing significant between-group differences in the plasma level ($P<0.05/177=2.8\times10^{-4}$, two-tailed, Bonferroni correction). Bars of metabolites within the same category were colored the same. **a)** Metabolites showing higher plasma levels in the existing depression compared to the prodromal depression group. **b)** Metabolites showing lower plasma levels in the existing depression compared to the prodromal depression group. A full list of abbreviations is provided in Supplementary Table 1.

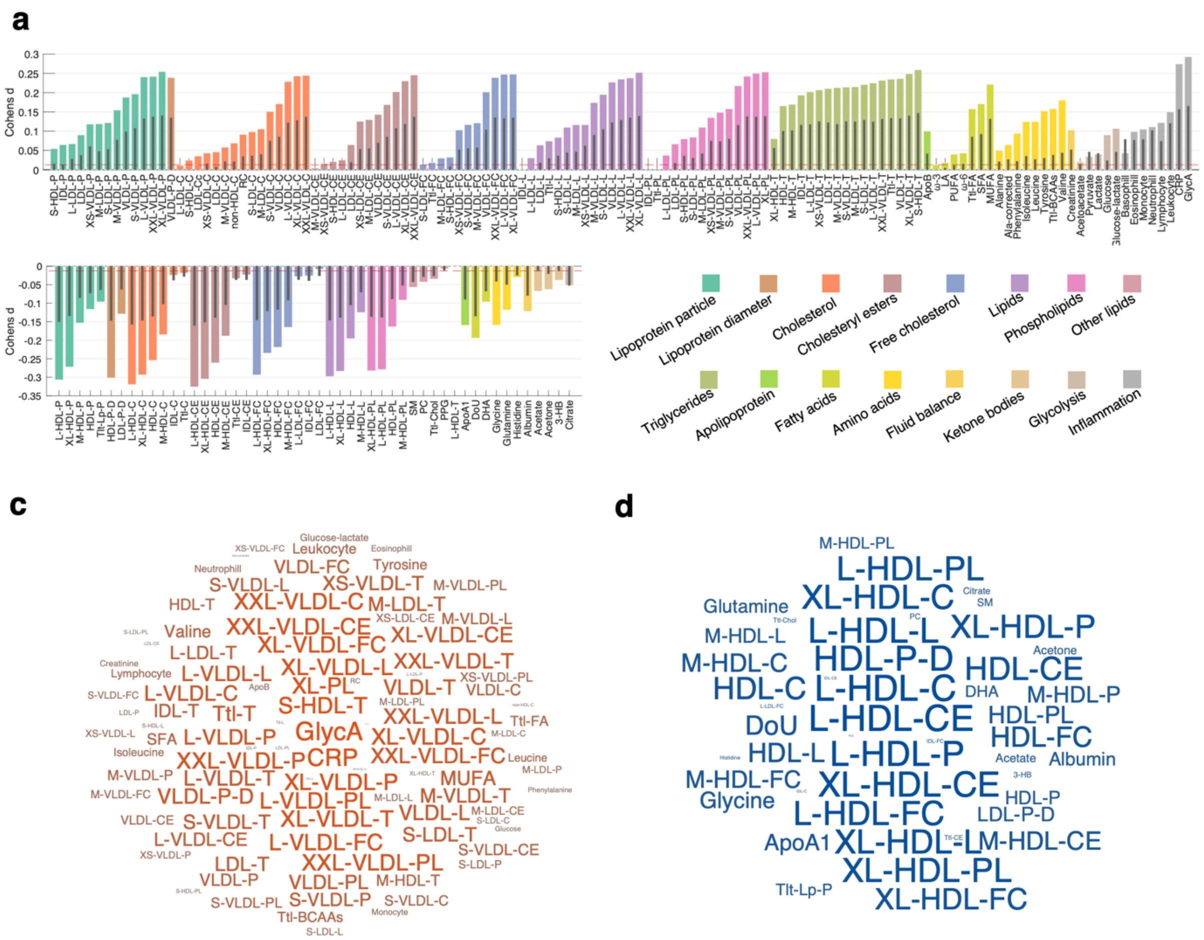


**Figure S3. Immunometabolic profile in metabolic disorders.** Bar plots show effect sizes (Cohen's $d$) for between-group differences in the plasma level of 177 metabolites across 16 metabolite categories. Coloured bars indicate differences between metabolic disorders and healthy individuals (HC), whereas the black lines inset indicate differences between existing depression and HC. Metabolite categories are shown with uniquely coloured bars. Dashed red line indicates the minimal effect size reaching statistical significance ($P$<0.05/177=2.8×10$^{-4}$, two-tailed, Bonferroni correction). **a)** Metabolites showing higher plasma levels in metabolic disorders compared to HC. **b)** Metabolites showing lower plasma levels in metabolic disorders compared to HC. **c,d)**. Word clouds show metabolites that significantly differ between metabolic disorders and HC. The font size is scaled according to the absolute value of the effect size, with red/blue indicates positive/negative Cohen's $d$ value respectively. A full list of abbreviations is provided in Supplementary Table 1.

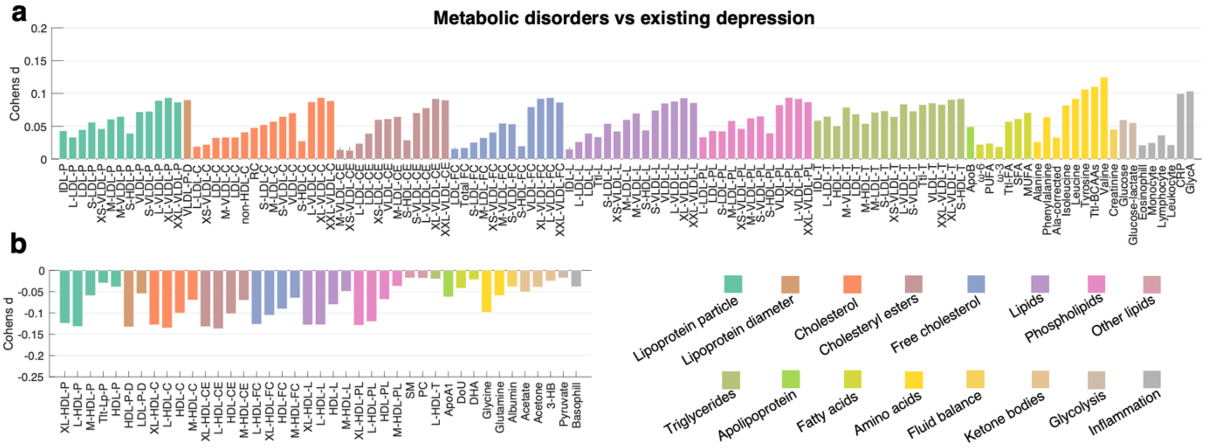


**Figure S4. Comparison of immunometabolic markers between metabolic disorders and existing depression groups.** Bar plots show effect sizes (Cohen’s $d$) of metabolites showing significant between-group differences in the plasma level ($P$<0.05/177=2.8×10$^{-4}$, two-tailed, Bonferroni correction). Bars of metabolites within the same category were colored the same. **a)** Metabolites showing higher plasma levels in metabolic disorders compared to existing depression. **b)** Metabolites showing lower plasma levels in metabolic disorders compared to existing depression. A full list of abbreviations is provided in Supplementary Table 1.

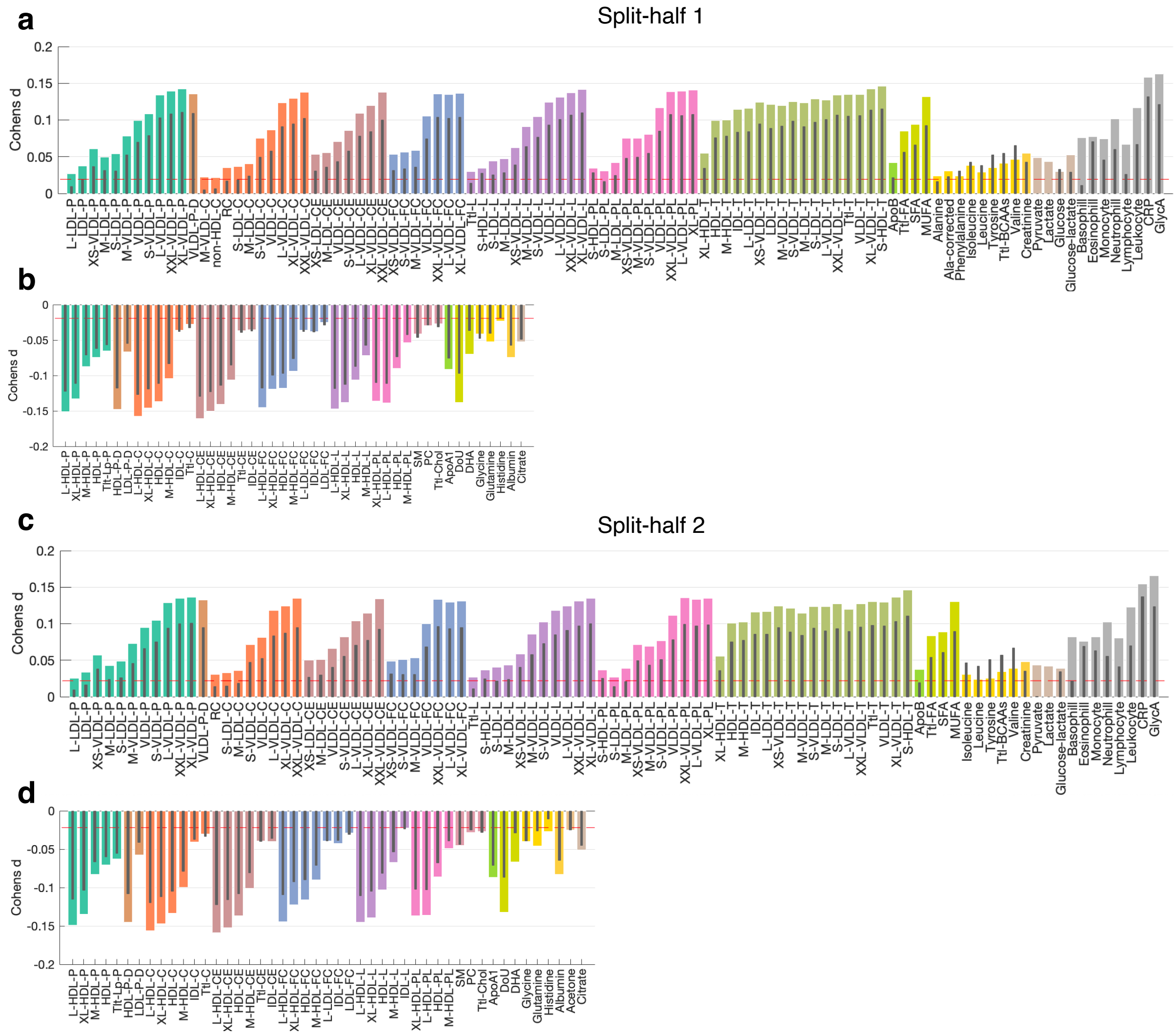


**Figure S5. Reproducible immunometabolic profiles in depression in split-half samples.** Bar plots show effect sizes (Cohen's $d$) for between-group differences in the plasma level of 177 metabolites across 16 metabolite categories. Coloured bars indicate differences between existing depression and healthy individuals, whereas the inset black lines indicate differences between prodromal depression and HC. Metabolite categories are shown with uniquely coloured bars. Dashed red line indicates the minimal effect size reaching statistical significance ($P<0.05/177=2.8\times10^{-4}$, two-tailed, Bonferroni correction). Non-significant metabolites are not shown. **a,b)** Metabolites showing higher (**a**) and lower (**b**) plasma levels in depression compared to HC in split-half 1 sample (existing: n=22,810; prodromal: n=12,210; HC: n=13,815) . **c,d)** Metabolites showing higher (**c**) and lower (**d**) plasma levels in depression compared to HC in split-half 2 sample (existing: n=22,957; prodromal: n=12,174; HC, n=13,821). A full list of abbreviations is provided in Supplementary Table 1.

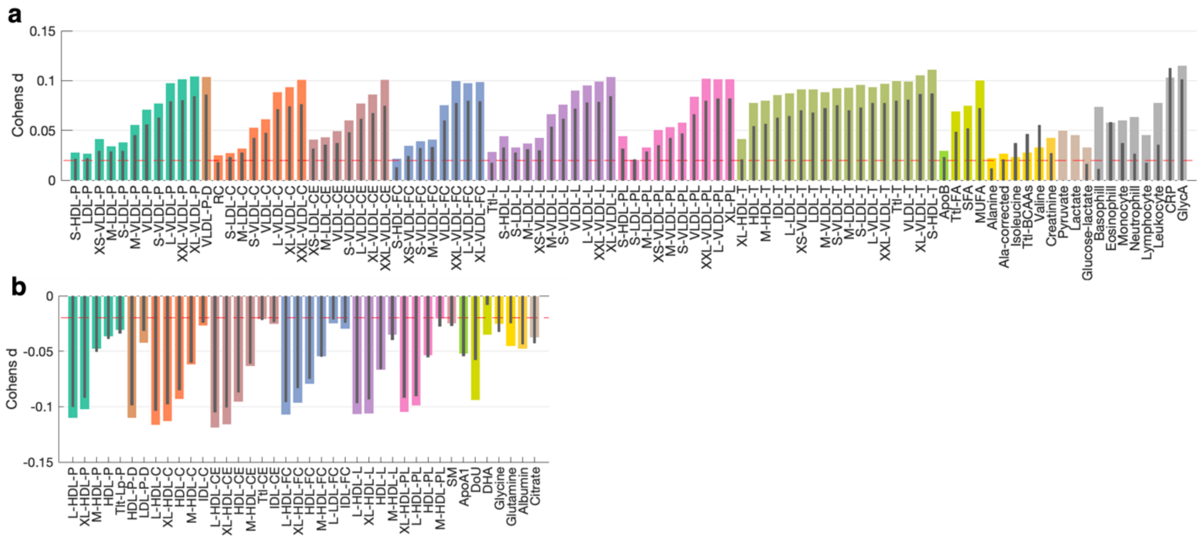


**Figure S6. Altered immunometabolic profiles in depression controlling for individual variation in socioeconomic status, lifestyle, early-life factors and genetic disposition for metabolic function.** Bar plots show effect sizes (Cohen's $d$) for between-group differences in the plasma level of 177 metabolites across 16 metabolite categories. Coloured bars indicate differences between existing depression and healthy individuals (HC), whereas the black lines inset indicate differences between prodromal depression and HC. Metabolite categories are shown with uniquely coloured bars. Dashed red line indicates the minimal effect size reaching statistical significance ($P$<0.05/177=2.8×$10^{-4}$, two-tailed, Bonferroni correction). Non-significant metabolites are not shown. **a)** Metabolites showing higher plasma levels in depression compared to HC. **b)** Metabolites showing lower plasma levels in depression compared to HC. Socioeconomic status: Townsend deprivation index, tertiary education; lifestyle: tobacco smoking, weekly alcohol consumption, usual walking pace, summed MET minutes per week; early-life factors breastfed as a baby, comparative body and height size at age 10, maternal smoking around birth; genetic disposition: polygenetic risk scores of BMI, HDL-cholesterol, LDL-cholesterol, and type 2 diabetes.

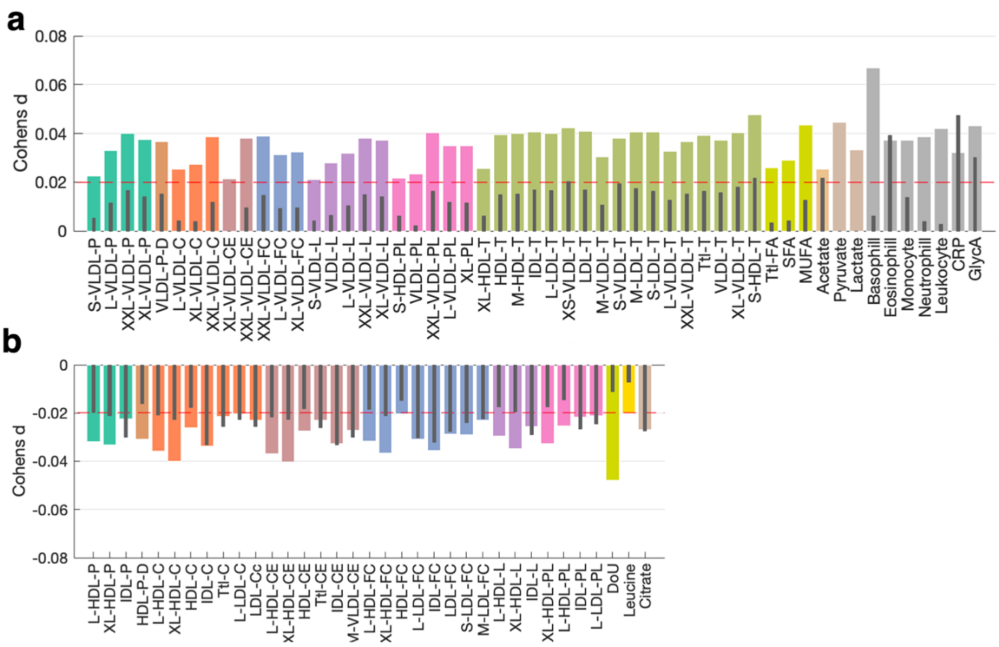


**Figure S7. Altered immunometabolic profiles in depression controlling for individual variation in BMI, socioeconomic status, lifestyle, early-life factors and genetic disposition for metabolic function.** Bar plots show effect sizes (Cohen's $d$) for between-group differences in the plasma level of 177 metabolites across 16 metabolite categories. Coloured bars indicate differences between existing depression and healthy individuals (HC), whereas the black lines inset indicate differences between prodromal depression and HC. Metabolite categories are shown with uniquely coloured bars. Dashed red line indicates the minimal effect size reaching statistical significance ($P<0.05/177=2.8\times10^{-4}$, two-tailed, Bonferroni correction). Non-significant metabolites are not shown. **a)** Metabolites showing higher plasma levels in depression compared to HC. **b)** Metabolites showing lower plasma levels in depression compared to HC. Socioeconomic status: Townsend deprivation index, tertiary education; lifestyle: tobacco smoking, weekly alcohol consumption, usual walking pace, summed MET minutes per week; early-life factors breastfed as a baby, comparative body and height size at age 10, maternal smoking around birth; genetic disposition: polygenetic risk scores of BMI, HDL-cholesterol, LDL-cholesterol, and type 2 diabetes.

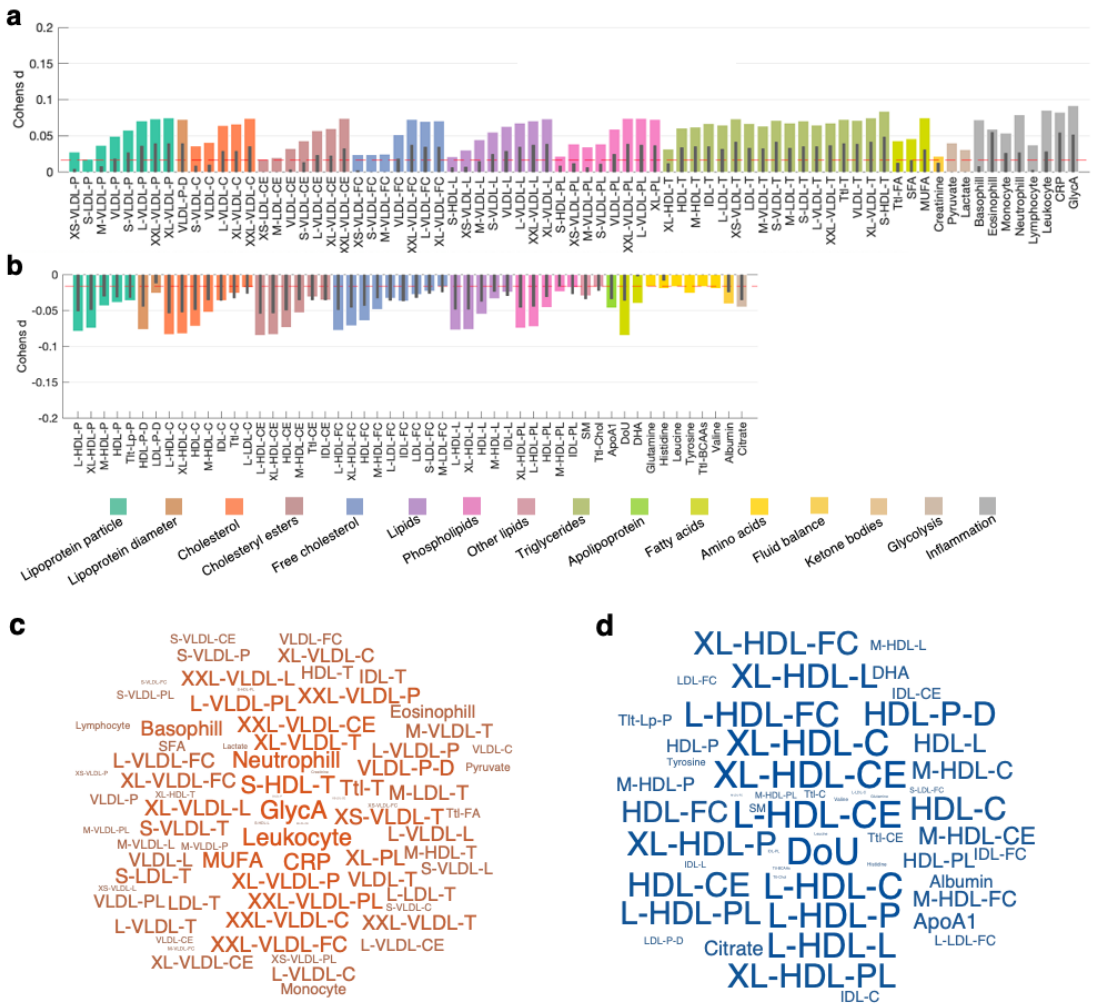


**Figure S8. Immunometabolic profiles in depression without established metabolic disorders.** Bar plots show effect sizes (Cohen's $d$) for between-group differences in the plasma level of 177 metabolites across 16 metabolite categories. Coloured bars indicate differences between existing depression (n=27,377) and healthy individuals (HC, n=27,636), whereas the inset black lines indicate differences between prodromal depression (n=16,084) and HC. Metabolite categories are shown with uniquely coloured bars. Dashed red line indicates the minimal effect size reaching statistical significance ($P<0.05/177=2.8\times10^{-4}$, two-tailed, Bonferroni correction). Non-significant metabolites are not shown**. a)** Metabolites showing higher plasma levels in depression compared to HC. **b)** Metabolites showing lower plasma levels in depression compared to HC. **c,d)**. Word clouds show metabolites that significantly differ between existing depression and HC. The font size is scaled according to the absolute value of the effect size, with red/blue indicating positive/negative Cohen's $d$ values respectively. A full list of abbreviations is provided in Supplementary Table 1.

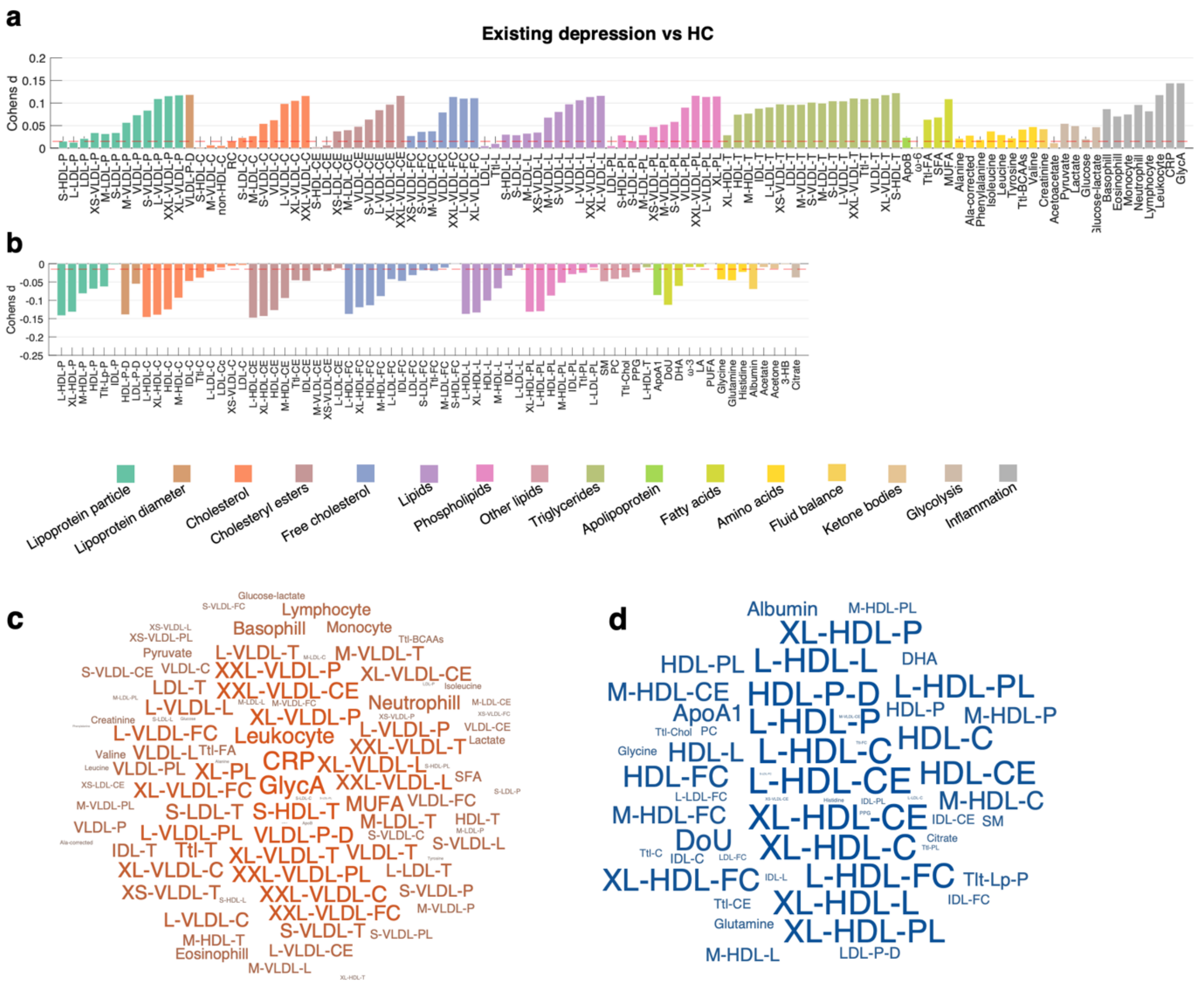


**Figure S9. Immunometabolic profiles in psychotropic-medication naïve depression.** Bar plots show effect sizes (Cohen's $d$) for between-group differences in the plasma level of 177 metabolites across 16 metabolite categories. Coloured bars indicate differences between existing depression (n=32,399) and healthy individuals (HC, n=27,636), whereas the black lines inset indicate differences between prodromal depression and HC. Metabolite categories are shown with uniquely coloured bars. Dashed red line indicates the minimal effect size reaching statistical significance ($P$<0.05/177=2.8×10$^{-4}$, two-tailed, Bonferroni correction). **a)** Metabolites showing higher plasma levels in depression compared to HC. **b)** Metabolites showing lower plasma levels in depression compared to HC. **c,d)**. Word clouds show metabolites that significantly differ between existing depression and HC. The font size is scaled according to the absolute value of the effect size, with red/blue indicates positive/negative Cohen's $d$ value respectively. A full list of abbreviations is provided in Supplementary Table 1.

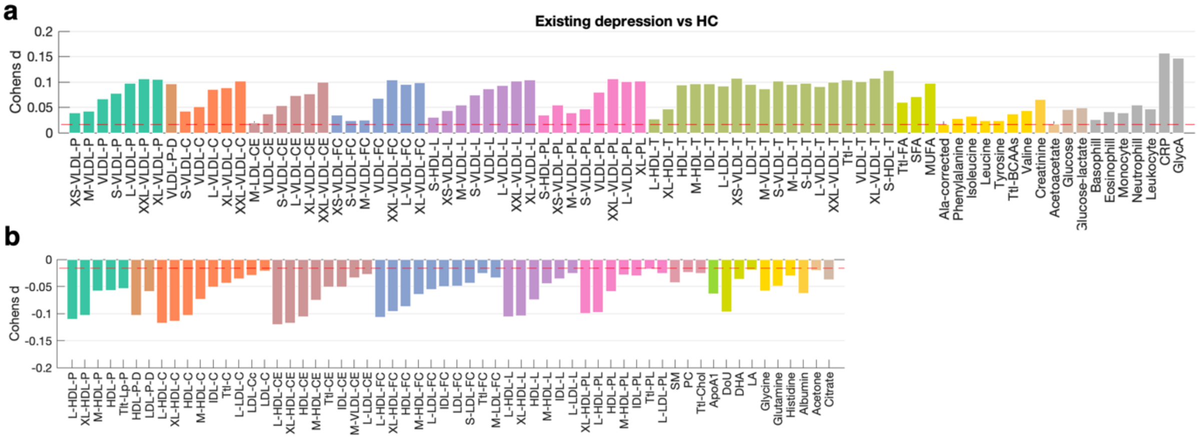


**Figure S10. Altered immunometabolic profiles in clinically diagnosed depression.** Bar plots show effect sizes (Cohen's $d$) for between-group differences in the plasma level of 177 metabolites across 16 metabolite categories. Coloured bars indicate differences between existing depression (n=23,707) and healthy individuals (HC, n=27,636). Metabolite categories are shown with uniquely coloured bars. Dashed red line indicates the minimal effect size reaching statistical significance ($P$<0.05/177=2.8×10$^{-4}$, two-tailed, Bonferroni correction). Non-significant metabolites are not shown. **a)** Metabolites showing higher plasma levels in depression compared to HC. **b)** Metabolites showing lower plasma levels in depression compared to HC. A full list of abbreviations is provided in Supplementary Table 1.

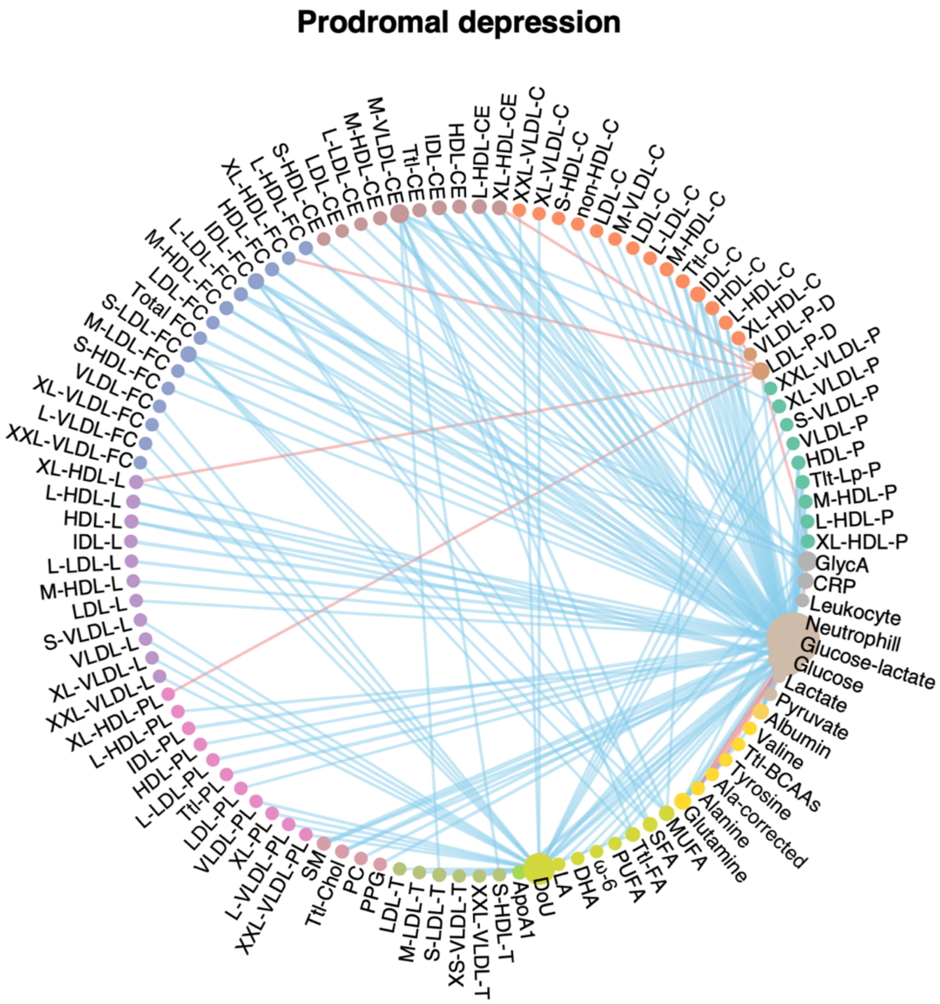


**Figure S11. Metabolomic networks in prodromal depression.** The strength of coupling between pairs of peripheral metabolites was compared between the depression and healthy comparison groups. Networks characterize metabolite pairs with significant between-group differences. Each network shows the top 1% of connections with significantly altered strength in prodromal depression (n=24,384) compared to healthy individuals (n=27,636, *P*<0.05, FDR corrected across 15,576 metabolite pairs). Bootstrapping (n=1,000) was used to estimate the distribution of the connectivity strength for each pair of metabolites in each group. In the graph, blue/red edge represents reduced/increased coupling between metabolites in depression compared to healthy individuals. Node size is modulated by the degree of nodal strength. Nodes of metabolites of the same category were colored the same. Edge thickness is modulated by the altered connectivity strength.

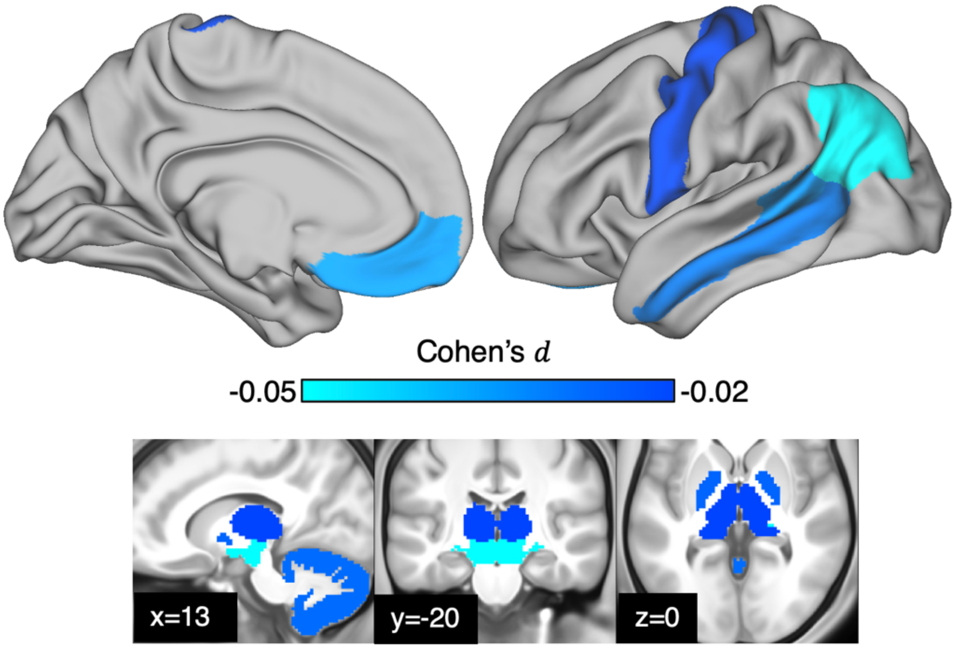


**Figure S12. Brain gray matter volume reduction associated with depression.** Effect sizes of brain regions that showed significantly reduced (*P*<0.05, uncorrected) gray matter volume in depression compared to healthy individuals are rendered on cortical surface (Desikan-Killiany atlas) and standard Montreal Neurological Institute (MNI)-152 anatomical space for visualization.
